# Differences in antibody kinetics and functionality between severe and mild SARS-CoV-2 infections

**DOI:** 10.1101/2020.06.09.20122036

**Authors:** Ger Rijkers, Jean-Luc Murk, Bas Wintermans, Bieke van Looy, Marcel van den Berge, Jacobien Veenemans, Joep Stohr, Chantal Reusken, Pieter van der Pol, Johan Reimerink

**Author notes:** Contributed equally to this work (Ger Rijkers, Jean-Luc Murk).

## Abstract

We determined and compared the humoral immune response in severe, hospitalized and mild, non-hospitalized COVID-19 patients. Severe patients (n=38) develop a robust antibody response to SARS-CoV-2, including IgG and IgA antibodies. The geometric mean 50% virus neutralization titer is 1:240. SARS-CoV-2 infected hospital personnel (n=24), who developed mild symptoms necessitating leave of absence, self-isolation, but not hospitalization, 75 % develop antibodies, but with low/absent virus neutralization (60% < 1:20).

While severe COVID-19 patients develop a strong antibody response, mild SARS-CoV-2 infections induce a modest antibody response. Long term monitoring will show whether these responses predict protection against future infections.

## Introduction

Severe acute respiratory syndrome virus 2 (SARS-CoV-2) which emerged in the human population at the end of 2019 had reached pandemic proportions by March 2020 [1,2]. The host defense against this new member of the corona virus family will depend on innate immunity, humoral and cellular immune responses, of yet unknown relative importance [3,4]. For these reasons, level of protection and longevity of protection cannot be established or predicted.

Similar to other respiratory viruses the primary diagnosis of COVID-19 is routinely made by RT-PCR detection of SARS-CoV-2. As the sensitivity of molecular detection relies on severity of illness, sample type and timing of sampling in the course of infection [5,6], serology has important additional value to establish a recent infection and to support patient care. It is therefore important to carefully determine the kinetics, magnitude and functionality of the humoral immune response in COVID-19 patients with different disease severity.

Here, we have analyzed the type and functionality of the humoral immune response of PCR-confirmed hospitalized COVID-19 patients, both ICU and non-ICU admitted, and of non-hospitalized patients with mild disease. We performed a semi-quantitative analysis of total Ig, IgG and IgA antibodies by ELISA as well as a functional analysis by virus neutralization assays.

## Methods

### Patients and blood sampling

Serum samples were collected from a prospective cohort of 38 RT-PCR-confirmed [7] COVID-19 patients (Table 1) admitted to the Admiral de Ruyter Hospital in Goes, The Netherlands in accordance with the local clinical procedures in the period March 2020 – May 2020. The presenting clinical symptoms included fever (n=17), coughing (18), dyspnea (11), dizziness and/or confusion (4) and general malaise (6). Patients were admitted to the hospital median 7 days (range 1–12) after onset of symptoms. 15 of 38 patients were admitted to the ICU. RT-PCR was always performed on the first hospital day on nasopharyngeal swabs. Serial blood sampling (3 times per week) was started a median 2 days (range 1–7) after positive RT-PCR.

**Table 1.** Patient characteristics

| Variable | Evaluable | Median (range)<br>/ n (%) | Intensive care |  | p-value |
| --- | --- | --- | --- | --- | --- |
|  |  |  | No (n =<br>23) | Yes (n =<br>15) |  |
| Age (years) | 38 | 70 (38 - 87) | 71 (38 - 87) | 68 (56 - 80) | 0.37<sup>1</sup> |
| Male gender | 38 | 26 (68) | 14 (61) | 12 (80) | 0.21<sup>2</sup> |
| Any comorbidities | 38 | 26 (68) | 13 (56) | 13 (87) | 0.11<sup>2</sup> |
| Diabetes mellitus | 38 | 4 (11) | 4 (17) | 0 (0) | 0.14<sup>2</sup> |
| Hypertension | 38 | 13 (34) | 8 (35) | 5 (33) | 0.79<sup>2</sup> |
| Coronary heart disease | 38 | 8 (21) | 6 (26) | 2 (13) | 0.59<sup>2</sup> |
| COPD | 38 | 10 (26) | 4 (17) | 6 (40) | 0.24<sup>2</sup> |
| Body Mass Index | 31 | 27 (19-41) | 28 (19-41) | 25 (22-27) | 0.14<sup>1</sup> |
| Mortality | 38 | 6 (16) | 4 (17) | 2 (13) | 0.90<sup>2</sup> |

<sup>1</sup> p-value of two-sided Student T-test
<sup>2</sup> p-value of Chi-square test
COPD: Chronic Obstructive Pulmonary Disease

The second cohort of 24 patients with mild COVID-19 disease consisted of hospital personnel (both from clinical departments as well as laboratory departments) who developed fever and/or coughing and/or dyspnea and were tested positive for SARS-CoV-2 by RT-PCR. They were asked to maintain self-quarantine until symptoms resolved. All of them were kept under control of their own GP and none of them required hospitalization.

The study was performed in accordance with the guidelines for sharing of patient data of observational scientific research in emergency situations as issued by the Commission on Codes of Conduct of the Foundation Federation of Dutch Medical Scientific Societies (https://www.federa.org/federa-english).

### SARS-CoV-2 immunoassays

The Wantai SARS-CoV-2 total antibody ELISA (Beijing Wantai Biological Pharmacy Enterprise, Beijing, China; Cat # WS1096) was performed according to the manufacturer’s instructions [8]. This assay is an double-antigen sandwich ELISA using the recombinant receptor binding domain antigen (RBD) of SARS-CoV-2 as antigen. SARS-CoV-2 IgG and IgA antibodies were determined by ELISA using the beta version of the EUROIMMUN immunoassay kit (EUROIMMUN Medizinische Labordiagnostika AG, https://www.euroimmun.com) according to the manufacturer’s protocol [8,9]. In this assay, wells are coated with recombinant structural protein (S1 domain) of SARS-CoV-2.

### Corona virus microarray

Sera were tested for the presence of IgG antibodies reactive with the four common human coronaviruses hCoV-OC43, hCoV-HKU1, hCoV-NL63 and hCoV-229E S1 subunit antigens in a protein microarray essentially as described before [10]. Antibody titers were defined as the interpolated serum concentration that provokes a response at 50% on a concentration-response curve between the minimum and maximum signal processed with R 2.12.1 statistical software as described before [11].

### SARS-CoV-2 Neutralization Assay

Sera were tested by a SARS-CoV-2-specific virus neutralization test (VNT) based on a protocol described previously with some modifications [12]. Briefly, serial dilutions of heat-inactivated samples (30 min 56°C) were incubated with 100 TCID50 of SARS-CoV-2 for 1h at 35°C. African green monkey (Vero-E6) cells were added in a concentration of 2 × 10^4^ cells per well and incubated for three days at 35°C in an incubator with 5% CO_2_. The VNT titer was defined as the reciprocal of the sample dilution that showed a 50% (VNT50) or 90% (VNT90) protection of virus growth. Samples with titers equal to 10 and higher were defined as SARS-CoV-2 seropositive.

### Statistical Analysis

Categorical variables were described as frequency rates and percentages, and continuous variables were described using geometric means, median, and interquartile range values. Means for continuous variables were compared using independent group t-tests. Categorical variables were compared using the Chi-square test. All analyses were done in Excel with the data analysis toolpack. P-values of less than 0.05 were considered statistically significant.

## Results

A prospective cohort of 38 consecutive hospitalized COVID-19 patients (Table 1) was monitored for development of anti-SARS-CoV-2 Ig, IgG and IgA antibodies. Within 2–5 days post onset of symptoms, 4 patients already showed total Ig responses and a steep increase in IgG ratio’s (see also below and Figure 1a–c). Most patients responded for IgG and IgA between day 10 and 15 (Figure 1b,c). The vast majority of the hospitalized patients developed high levels of antibodies after 3–4 weeks: 100% of patients had detectable (total)antibodies, 84% had detectable IgG antibodies, and 92% IgA antibodies.

**Figure 1.**
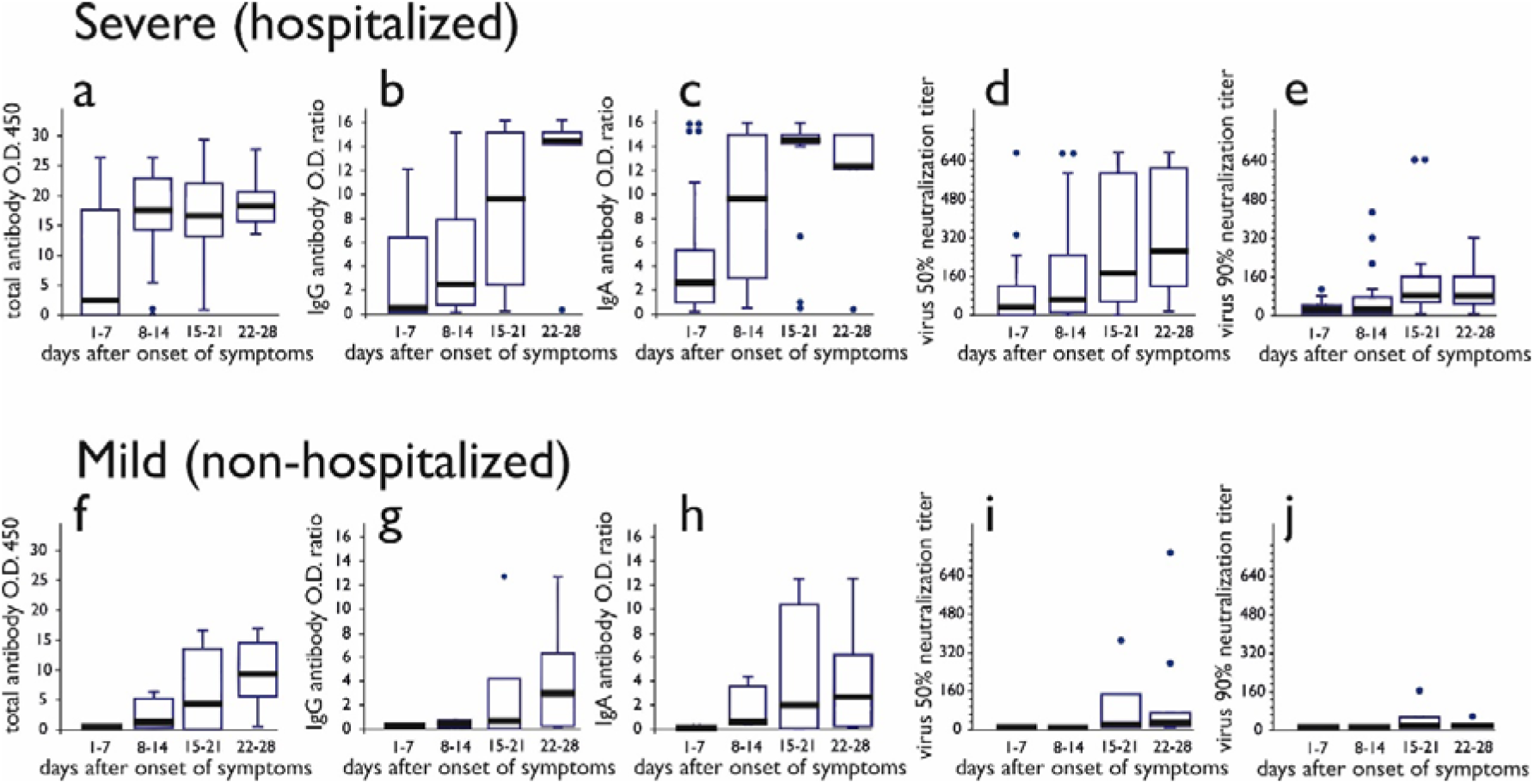
Quantitative and qualitative antibody response against SARS-CoV-2 in severe (panels a-e) and mild COVID-19 patients (panels f-j). Shown are total (panels a and f), IgG (panels b and g), IgA (panels c and h) antibody levels as well as virus neutralization titer at 50% neutralization (panels d and i) and 90% neutralization (e and j). The horizontal line in the middle of each box indicates the geometric mean, the top and bottom borders of the box mark the 75th and 25th percentiles, the whiskers above and below the box indicate the range. Outliers, with values > 1.5 the interquartile range, are indicated with individual dots.

Patients admitted to the ICU (n= 15) showed an antibody response similar to patients in the general COVID-19 ward (n=23), with an IgG antibody ratio at day 14–21 of 9.9 ± 5.2 (ICU) and 8.7 ±5.3 (ward), p>0.05; IgA antibody ratio was ≥ 15 (ICU) and 13.6 ± 2.8 (ward), p>0.05).

The functionality of the antibodies in the hospitalized cohort was assessed by VNT50 and VNT90 (Figure 1, panels d and e). In the first week upon onset of symptoms, 43% of the patients had detectable VNT50 (median 22, range10 to 640). At 21–28 days all patients had detectable neutralizing antibody titers in the VNT50 with a median titer of 226 (range 20 to 800). Median titer in the VNT90 was 50 (range <10–240).

As indicated above, 4 patients already showed high IgG levels (O.D. ratio > 5) and even plateau levels of IgA antibodies early during the course of disease, i.e. within 10 days after onset of symptoms. To exclude cross-reactivity due to (recent) previous infection with one of the four common coronaviruses (i.e hCoV-OC43, hCoV-NL63, hCoV-HKU1 and hCoV-229E) in these so called “early IgA responders” antibodies titers against these common CoVs were determined in two early responders (Figure 2a,b) and compared with those in two other COVID-19 patients (Figure 2c,d). We found minimal and inconsistent fluctuations in IgG against the four common CoV across all four patients indicating that the early IgA and total Ig anti-SARS-CoV-2 responses for some patients were not the result of cross-reactivity due to a previous common CoV infection.

**Figure 2.**
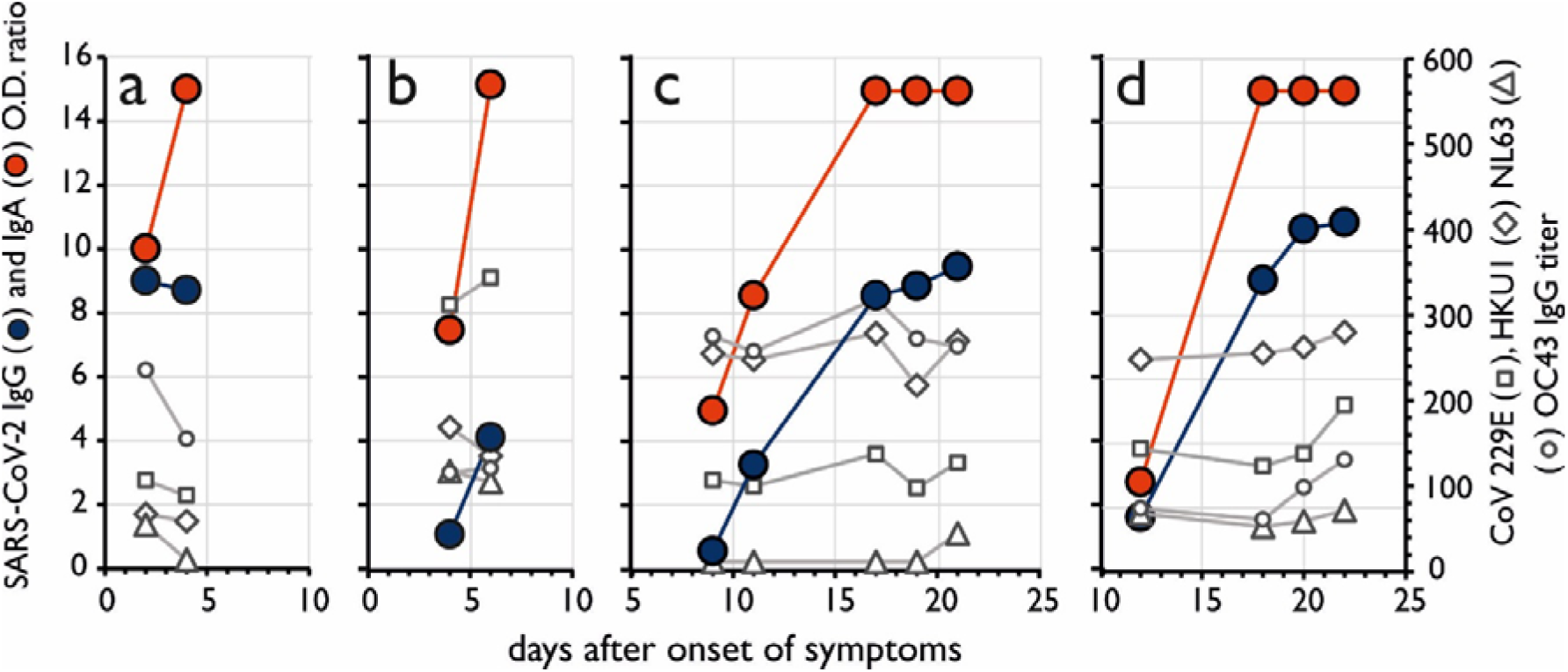
Antibody levels against circulating corona viruses during the course of COVID-19 disease. Each panel (a-d) represents a single patient. IgG and IgA SARS-CoV-2 antibodies displayed by blue and red circles, respectively, and expressed as O.D. ratio values on the left Y-axis. hCoV 229E (□), HKU1(◊), NL63 (△), and OC43 (О) IgG antibody titers are shown as open symbols and expressed as titers.

Next, we investigated the antibody responses in a cohort of hospital personnel who had tested positive in the SARS-CoV-2 RT-PCR but had only experienced mild clinical symptoms. 87% developed SARS-CoV-2 total antibodies by day 21–28, with a significant lower titer than severe patients (GMT 4.9 and 17.3, respectively, p < 0.001). The median titer in the VNT50 was 29 (range <10 to 640) and in VNT90 was 10 (range <10–12), both significantly lower than in severe patients (p values < 0.0001) (Figure 1, panels d and e vs i and j). 33% of the mild patients remained negative for the presence of virus neutralizing antibodies.

## Discussion

Serial blood sampling of 38 severe (hospitalized) and 24 mild COVID-19 patients allowed for detailed analysis of the kinetics and magnitude of the SARS-CoV-2 antibody response in patients with different levels of disease severity. At 2–4 weeks after onset of symptoms detectable total, IgG and IgA antibodies were found in 100%, 86% and 94% of the severe patients, and 81%, 81%, 61% of the mild patients respectively. Virus neutralizing activity was demonstrable in the vast majority of severe patients (all at VNT50, 95% at VNT90) but only in 65% (VNT50) and 30% (VNT90) in the mild patients. We did not find a significant difference in the kinetics, magnitude, nor functionality of the response between hospitalized patients of different disease severity (routine ward vs intensive care vs fatal outcome). This is in accordance with findings in hospitalized SARS-CoV-1 patients [13]. It is however possible that larger series of hospitalized patients with variable clinical outcome can reveal differences in the nature and kinetics of the immune response, because our sample size was limited.

Of 2 of our patients we had serum samples available from the period before onset of COVID-19. As expected, total antibody, IgG and IgA anti-SARS-CoV-2 antibodies were not demonstrable at that time.

Early responders (especially levels of IgG and IgA within 5 days after onset of symptoms) were only observed in the hospitalized cohort and could have been due to a prolonged pre-symptomatic period, or recall bias with respect to onset of complaints [14]. Antibody levels against hCoV-OC43 and hCoV-HKUI in early responder patients did not differ from other patients (data not shown) and antibody levels against circulating coronaviruses did not change during the course of COVID-19 in both very early and “normal” responders while SARS-CoV-2 directed IgA and IgG levels did (Figure 2).

Two of our patients with severe disease who survived failed to show an IgG response at day 21 after disease onset (although the total antibody assay was positive). Our two IgG negative patients, 69 and 87 years of age, had persistently positive PCR test results on respectively day 28 and day 37 after disease onset. Patients with an inadequate IgG antibody response may exhibit prolonged viral shedding, and thus longer periods of infectivity. Prolonged viral RNA shedding has been reported previously [15], and further studies that include viral cultures are needed to investigate the prolonged infectivity hypothesis. One (other) patient remained negative for IgA antibodies, although this patient, as well as all others, had normal serum IgA immunoglobulin levels (data not shown). Severe COVID-19 patients remaining seronegative have also been reported by others (e.g. [6,8]). It can be speculated that their antibody response is dominated by epitopes not represented in the immunoassays we have used. Antibody testing against a more extensive array of SARS-CoV-2 peptides will be required to address this possibility.

From our data it is clear that hospitalized COVID-19 patients mount a robust humoral immune response against SARS-CoV-2, including antibodies with virus neutralizing activity. Mild infections with SARS-CoV-2, with clinical symptoms not requiring hospital admission, do show an antibody response but delayed in comparison to severe patients and with minimal functional activity. Long term monitoring will be required to determine whether these quantitative and qualitative parameters of the humoral immune response predict protection against future infection.

## Data Availability

not applicable

## Acknowledgements

We would like to thank all the personnel in the laboratory to make this evaluation possible. Especially we would like to thank (in alphabetical order): Fion Brouwer, Gert-Jan Godeke, Gabriel Goderski, Marieke Hoogerwerf, and Ilse Zutt.

## Notes

### Competing Interest Statement

The authors have declared no competing interest.

### Funding Statement

No external funding was received

### Author Declarations

Exemption for the research was provided by Medisch ethische toetsingscommissie Brabant (Medical Ethical Committee Brabant) PO Box 90151, 5000 LC Tilburg The Netherlands info@metcbrabantnl

